# Higher loss of livelihood and impoverishment in households affected by tuberculosis compared to non-tuberculosis affected households in Zimbabwe: a cross-sectional study

**DOI:** 10.1101/2023.12.05.23299470

**Authors:** Collins Timire, Rein MGJ Houben, Debora Pedrazzoli, Rashida Abbas Ferrand, Claire J Calderwood, Virginia Bond, Fredrick Mbiba, Katharina Kranzer

## Abstract

**Introduction:** Tuberculosis (TB) disproportionally affects poor people, leading to income and non-income losses. Measures of socioeconomic impact of TB, e.g. impoverishment and patient costs are inadequate to capture non-income losses. We applied impoverishment and a multidimensional measure on TB and non-TB affected households in Zimbabwe.

**Methods:** We conducted a cross-sectional study in 270 households: 90 non-TB; 90 drug-susceptible TB (DS-TB), 90 drug-resistant TB (DR-TB) during the COVID-19 pandemic (2020-2021). Household data included ownership of assets, number of household members, income and indicators on five capital assets: financial, human, social, natural and physical. We determined proportions of impoverished households for periods 12 months prior and at the time of the interview. Households with incomes below US$1.90/day were considered to be impoverished. We used principal component analysis on five capital asset indicators to create a binary outcome variable indicating loss of livelihood. Log-binomial regression was used to determine associations between loss of livelihood and type of household.

**Results:** TB-affected households reported higher previous episodes of TB and household members requiring care than non-TB households. Households that were impoverished 12 months prior to the study were: 21 non-TB (23%); 40 DS-TB (45%); 37 DR-TB (41%). The proportions increased to 81%, 88% and 94%, respectively by the time of interview. Overall, 56% (152/270) of households sold assets: 44% (40/90) non-TB, 58% (52/90) DS-TB and 67% (60/90) DR-TB. Children’s education was affected in 31% (56/180) of TB-affected compared to 13% (12/90) non-TB households. Overall, 133(50%) households experienced loss of livelihood, with TB-affected households twice as likely to experience loss of livelihood; adjusted prevalence ratio (aPR=2.02 (95%CI:1.35-3.03)). The effect of TB on livelihood was most pronounced in poorest households (aPR=2.64, (95%CI:1.29-5.41)).

**Conclusions:** TB-affected households experienced greater socioeconomic losses compared to non-TB households. Multidimensional measures of TB are crucial to inform multisectoral approaches to mitigate impacts of TB and other shocks.

## Introduction

Tuberculosis (TB) disproportionally affects socioeconomically deprived people and leads to income and non-income losses.[1] While TB diagnostic tests and treatment services are usually provided free-of-charge, other medical costs such as hospitalisations, radiology services and blood tests are often not covered.[2] Households also incur social costs (stigma, social exclusion, deterioration of relations with neighbours and landlords) and substantial non-medical costs related to travel, accommodation and food, in addition to loss of income before, during and even post-treatment.[3]–[8] This, coinciding with reductions in household income, leads to severe socioeconomic burden.[9]

The impact of TB on households is even more pronounced in the context of drug resistant TB (DR-TB).[1] DR-TB treatment is longer than treatment for drug susceptible TB (DS-TB). Historically, people with DR-TB have been hospitalised (e.g. for injectable medications), and often experienced severe disease (in part due to treatment delays), complications and post-TB disability. Treatment delays may result from delayed health seeking or people being incorrectly started on DS-TB regimens before drug resistance is identified and people are switched to effective regimens.[10] Consequently, people with DR-TB experienced more direct and indirect costs than people with DS-TB.

Estimating the socioeconomic impact of TB on households is fraught with challenges. The most commonly employed measures of impact of TB are impoverishment and/or patient costs. The former determines the proportion of households that are pushed further into poverty by TB by comparing per capita income per day against a threshold, usually the international poverty line of United States Dollar (US$) 1.90 per person per day.[11] Patient costs surveys (PCS) broadened the scope for measuring impact of TB on households by collecting data on direct medical (consultations, ancillary medicines), direct non-medical (food, transport) and indirect costs (income loss).[12], [13] If the fraction of total costs (direct medical, direct non-medical and indirect costs) that are incurred by households as a fraction of a household’s annual income prior to TB exceeds 20%, the household is considered to have experienced catastrophic costs.[14] Global estimates of catastrophic costs, as measured through nationally representative PCS have revealed higher pooled prevalence of catastrophic costs in DR-TB affected households (82%) compared to DS-TB affected households (39%).[15]–[19] While PCS have provided valuable data on the socioeconomic impact of TB, their outcome measure (catastrophic costs) is benchmarked against income. This may lead to overestimation of the impact of TB among poor people who are likely to have unstable employment and income.

TB affects all facets of human wellbeing, leading to both income and non-income related losses. The sustainable livelihood framework (SLF),[20] is a useful framework to inform multidimensional and holistic estimates of socioeconomic impacts of TB. The framework conceptualises that households live in a vulnerability context characterised by various shocks, such as TB, and they make use of five available capital assets (human, financial, social, physical and natural capital) and various livelihood (coping) strategies in order to mitigate impacts of such shocks.[20] The livelihood strategies are either accumulative (households amassing assets in times of plenty to create buffers against future shocks) or coping (survival) strategies in order to survive shocks.[21]–[23] The coping strategies may be either harmful or non-harmful to livelihoods. Households may adopt short-term, non-harmful coping strategies e.g. spending savings, borrowing.[24] However, prolonged and/or sudden shocks may force households to expend resources rapidly and to adopt harmful coping strategies e.g. borrowing at exploitative interest rates and sale of productive assets.[24]–[26] The coping strategies that are adopted determine the five capital assets that are available in households, and whether households are resilient or vulnerable to shocks.[26]

Quantitative measures based on the SLF have been used to study the impact of shocks on household livelihoods in the context of agroforestry and climate change.[20], [27], [28] A similar approach could be used to measure the impact of TB.[29]

This study applies impoverishment and a SLF-based measure we previously proposed,[29] to assess the socioeconomic impact of TB on households overall, and stratified by DR-TB or DS-TB, compared to non-TB affected households in the same communities.

## Methods

### Study Setting

This study was conducted in four provinces of Zimbabwe: Harare and Bulawayo (both predominantly urban), Masvingo (urban and rural) and Matabeleland South (predominantly rural) (**Supplementary Figure 1)**. Zimbabwe, a southern African country, had an estimated TB and DR-TB incidence of 190/100,000 population and 4.9/100,000 population in 2021.[30], [31] Treatment success (completion and cure) was 83% for people with DS-TB and 54% among people with DR-TB.[32]–[34] The prevalence of TB/HIV co-infection was 50%. The prevalence of HIV in the general adult population is estimated at 12.9%, but is much higher (17.6%) in Matabeleland South province.[35] Zimbabwe has experienced socioeconomic challenges in the past two decades, with unemployment as high as 90% in 2015.[36] In 2020, the Human Development Index was only 0.571, placing it 150^th^ out of 189 countries.[37] About 72% of Zimbabwean population live below the poverty line of US$1.90 per day.[36]

This study was conducted during the COVID-19 pandemic (October 2020-March 2021) and as a result there were several COVID-19 waves and various degrees of national lockdowns during the study period. Zimbabwe recorded the first case of COVID-19 in March 2020, resulting in lockdowns where businesses were shut and health workers were reassigned to COVID-19 related work.[38], [39] The government’s social protection scheme, the Harmonised Social Cash Transfer, initially meant for food insecure households,[40] was activated to cushion vulnerable households during lockdowns. The highest disbursement was US$25 per household per month. However this was converted to local currency equivalent at prevailing interbank rates which are often much lower than the black market rates on which most retail operates. Consequently, the US$25 disbursement was in fact worth very little: enough to buy five kilogrammes of maize flour which could feed a household of five people with their staple carbohydrate for at most a week.

### Management of TB in Zimbabwe

TB treatment in Zimbabwe is decentralised to primary health facilities. TB molecular diagnostics, e.g Xpert MTB/Rif assay (Cepheid, Sunnyvale, CA, USA), and treatment are provided free-of-charge. However, costs incurred prior to diagnosis, including clinic fees, hospitalisation costs, radiology investigations and laboratory tests are not covered. Radiology and many laboratory tests are mostly unavailable in public facilities and are usually accessed from private health providers, resulting in significant out-of-pocket costs. An all-oral 9-month DR-TB treatment regimen was introduced in 2021, replacing the longer 18-24 month injection-based regimen.[41] People on DR-TB treatment are eligible for non-contributory social protection in the form of conditional cash transfers (CCTs). Once registered, they receive US$25 per month till treatment completion, death or loss-to-follow up, whichever comes first. However, the cash transfer is subject to delays, unpredictable disbursements and has modest coverage.[42]

### Study design and population

In this cross-sectional study, adults (≥18 years) who were alive and on treatment for DR-TB and DS-TB at 35 selected health facilities **(Supplementary Figure 1)** during the study period were eligible for inclusion. Health facilities were selected based on DR-TB caseloads in 2018. For each person with DR-TB, an age (within 5 year age-bands) and sex-matched person with DS-TB was identified from the same health facility’s TB register.

In each facility, data, including age, sex, date of treatment initiation, treatment regimen and mobile phone numbers were extracted from TB registers. Prospective participants were called by the study team who briefly described the purpose and procedures of the study and how the contact details for prospective participants had been obtained. Face-to-face meetings were arranged with those who expressed interest in the study. Those who were willing to participate were asked for written informed consent.

Community control households within 500 metres of the DR-TB affected households were selected. Hereafter, these households are referred to as ‘non-TB households’. These people and their households were also matched for age and sex with those affected by DR-TB and DS-TB in the ratio 1:1:1. To protect confidentiality, five households neighbouring DR-TB affected households on the same street in either direction were not approached for participation.

### Data collection

Data were collected using interviewer administered paper-based individual and household questionnaires. If the person with TB was the head of household, a household questionnaire was also administered, otherwise consent was sought from the head of household to administer a household questionnaire. Individual questionnaires captured socioeconomic details, experiences of stigma, duration from onset of TB symptoms to diagnosis of TB, past medical history, money spent on travel and medical expenses, income 12 months before the interview, income at time of interview, receipt of any social protection (only for people with DR-TB), any relocation and physical fitness. Stigma was measured using the scale adapted by Marangu et al.[43] The scale has 13 items capturing internalised stigma (4 items), perceived stigma (4 items) and general stigma (5 items). Each item was measured on a Likert scale ranging from 0-4: 0 indicating “Never”; 1 “Rarely”; 2 “Occasionally”; 3 “Regularly” and 4 indicating “Always”. Household questionnaires included questions on household asset ownership, current and past income, size of household, sale of assets (including selling and market prices of assets), borrowing, failure to repay loans, pledging crops or cattle, spending savings, withdrawal of children from school or transfers to cheaper schools, changes in social relations, changes of head of household and deaths in the household. The variables in the household questionnaire were informed by the SLF,[20] variables in patient cost surveys,[44] and indicators adapted from a study investigating livelihood in the context of HIV in Zimbabwe.[45] The SLF indicators are presented in **Supplementary Table 1**. All interviews were held in private locations suggested by participants.

### Data analysis

Data were entered in EpiData v3.1 (EpiData Association, Odense, Denmark) and were exported to Stata version 13 (StataCorp, College Station, TX, USA) for cleaning and analysis. The exposure of interest was type of household: non-TB affected or TB affected household. TB affected households were further categorised into DS-TB and DR-TB households. Categorical variables were summarised using frequencies and proportions. Differences in proportions were compared using the chi-square test. Continuous variables were summarised using medians and interquartile ranges (IQRs) and differences were compared using the Mann-Whitney-U test. Some variables were derived during analysis. These include effect on education of children (which was a composite of withdrawal of children from school or transferring children to cheaper schools or both); loss of household income (binary variable (yes/no) was determined when income at time of study was lower than income 12 months before the interview); dissavings (sale of assets, spending savings, taking loans); and changes in social relations comparing 12 months before and at the time of the interview based on self-reports by participants using Likert scales ranging from 1-10. A reduction in score was reflective of deteriorating social relations. To calculate impoverishment, we divided monthly household income by 30.5 and by the number of household members to determine income per person per day and classified households as impoverished when income per capita was below the poverty line of US$1.90 per day.[46] Monetary values reported in South African Rand (commonly used in Masvingo and Matabeleland South provinces) were converted to US$ for calculations using the Oanda currency converter (http://www.oanda.com). Total stigma was calculated by averaging the scores for the 13 scale items and individuals were considered to have experienced stigma when the average score was ≥2. We calculated mean scores for each of the five capital indicators and presented results as spider diagrams. We used principal component analysis (PCA) to categorise households into tertiles (poorest, poor and not-so-poor) based on household asset ownership and to reduce data on the five capital assets and coping strategies into a dichotomous outcome variable indicating loss of livelihood as previously described.[29] Details of the PCA are described in the **Supplement file.** Log binomial regression was used to test associations between loss of livelihood and type of household. We adjusted the analysis for previous TB episode (an indicator of previous shock for a household) in a multivariable Poisson regression, excluding matching variables (age, sex and province), and presented results as prevalence ratios (PRs) and adjusted PRs.

### Ethics

Ethical approval was obtained from the London School of Hygiene & Tropical Medicine Research Ethics Committee (22579), the Biomedical Research and Training Institute Institutional Review Board (AP160/2020) and the Medical Research Council Zimbabwe (MCRZ/A/2645).

## Results

We approached 285 people, of whom 270 (95%) people (and all corresponding heads of households) consented to take part in the study. Non-TB affected participants were less likely to be living with HIV, to report hospitalisations and to have relocated 12 months prior the study compared to TB-affected participants. People on DR-TB treatment incurred 2.7 times higher TB-related costs than people on DS-TB treatment (p<0.001). Of the 62/90 people with DR-TB who registered for CCTs, only 40 (65%) reported receiving any cash disbursement. TB stigma was experienced by 22 (24%) of people with DR-TB compared to 12 (13%) people with DS-TB (p=0.06) (**Table 1).**

**Table 1:** Baseline characteristics of participants (individual-level questionnaire) who were enrolled in the study.

| Characteristic |  | Non-TB<br>N=90<br>n (%) | DS-TB<br>N=90<br>n (%) | DR-TB<br>N=90<br>n (%) | p-value |
| --- | --- | --- | --- | --- | --- |
| Men |  | 50 (56%) | 50 (56%) | 50 (56%) | 1.00 |
| Age category | ≤24 | 7 (8%) | 12 (13%) | 12 (13%) | 0.64 |
|  | 25-34 | 27 (30%) | 30 (33%) | 24 (27%) |  |
|  | 35-44 | 33 (37%) | 33 (37%) | 36 (40%) |  |
|  | 45-54 | 14 (16%) | 12 (13%) | 10 (12%) |  |
|  | 55+ | 9 (10%) | 3 (3%) | 7 (8%) |  |
| Education of person | Primary | 8 (9%) | 17 (19%) | 25 (28%) | 0.03 |
|  | Secondary | 72 (80%) | 66 (73%) | 59 (65%) |  |
|  | Tertiary | 10 (11%) | 7 (8%) | 6 (7%) |  |
| HIV<sup>1</sup> positive |  | 27 (30%) | 58 (64%) | 60 (67%) | <0.001 |
| History of TB |  | 14 (16%) | 10 (11%) | 26 (29%) | 0.01 |
| Phase of TB treatment | Intensive | N/A | 51 (57%) | 17 (19%) | <0.001 |
|  | Continuation | N/A | 39 (43%) | 73 (81%) |  |
| Experienced TB stigma | Yes | N/A | 12 (13%) | 22 (24%) | 0.06 |
| Interval from symptoms to diagnosis (weeks) |  | N/A | 9 (6-22) | 13 (5-24) | 0.29 |
| Hospitalization in the past 12 months |  | 2 (2%) | 18 (20%) | 30 (41%) | <0.001 |
| Loss of income in the past 12 months |  | 46 (51%) | 74 (82%) | 79 (88%) | <0.001 |
| Changed residency in the past 12 months |  | 8 (9%) | 29 (32%) | 33 (37%) | <0.001 |
| TB related costs (USD) | Median (IQR) | N/A | 150 (100-275) | 400 (244-728) | <0.001 |

SES=Socioeconomic status; HH=Household; DS-TB=drug susceptible tuberculosis; DR-TB=drug resistant tuberculosis; IQR = interquartile range; USD = United States Dollar, <sup>1</sup> 1=unknown

Across the three strata, households were similar with respect to sex of head of household, socioeconomic status, education of head of household and the percentage having experienced a death in the household over the past 12 months. A higher proportion of TB-affected households had experienced TB before and reported that a household member had needed to be taken care of 12 months prior to the interview, compared to non-TB households (**Table 2).**

**Table 2:**
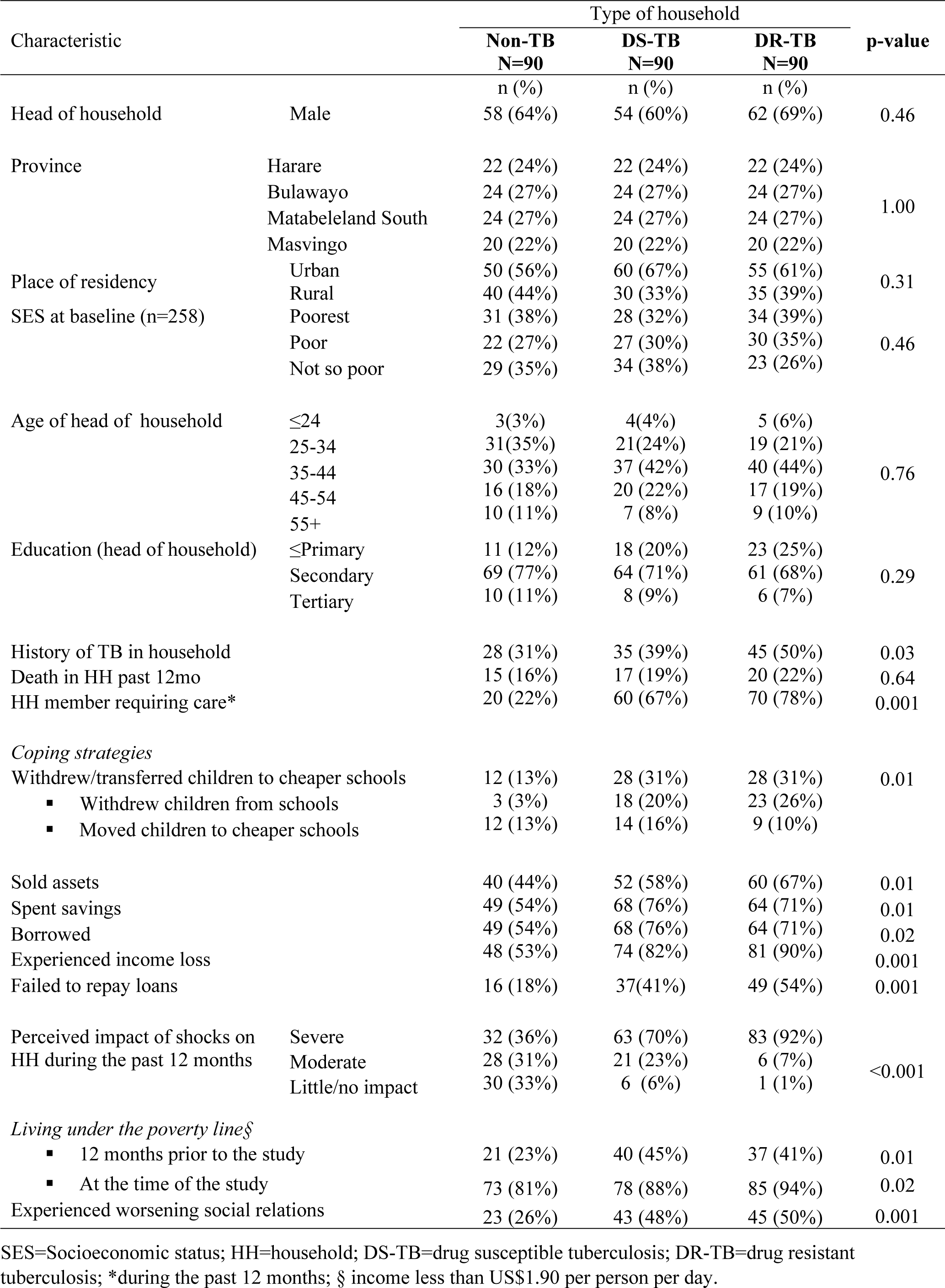
Baseline characteristics of study households that were enrolled in the study.

Twelve months prior to the study, the number (percentage) of non-TB, DS-TB and DR-TB households that were impoverished was 21 (23%), 40 (45%) and 37 (41%) (p=0.01). At the time of interview, the proportions increased to 81%, 88% and 94%, respectively (p=0.02).

TB-affected households experienced higher dissavings (borrowing, selling assets, spending savings) as compared to community households **(Table 2**). Overall, 56% (152/270) of households sold assets: 44% (40/90), 58% (52/90), 67% (60/90) of non-TB, DS-TB and DR-TB households, respectively (p=0.01). Median TB-related costs were higher among DR-TB households compared to DS-TB and non-TB households (DR-TB: US$400 [IQR:244-728] vs DS-TB: US$150 [IQR:100-275], p<0.001; **Table 2)** Almost a third of TB-affected households (n=56, 31%) reported that the education of children was negatively affected compared to one in eight (n=12, 13%) of non-TB households. Heads of households in 83 (92%) DR-TB and 63 (70%) DS-TB households reported their livelihoods were severely affected in the past 12 months compared to 32 (36%) heads of non-TB households.

Huge impacts on financial, human and social capitals were experienced in TB-affected compared to non-TB households (**Figure 1).** Overall, 133 (50% [95% confidence interval (CI): 44%-56%]) of households experienced loss of livelihood. Loss of livelihood was higher in DR-TB (62%) and DS-TB (60%) affected households compared to non-TB households (27%). TB affected households were two times more likely to experience loss of livelihood as compared to non-TB households, after adjustments for socioeconomic tercile and history of TB in the household (adjusted prevalence ratio [aPR]=2.02 [95%CI: 1.35-3.03]). There were no differences in loss of livelihood comparing DR-TB and DS-TB households (aPR=0.99, 95%CI:0.79-1.24; **Table 3).** The proportion of households experiencing loss of livelihood was 60% in the poorest households compared to 33% in the not-so-poor households (**Table 3**).The effect of TB on loss of livelihood was worst in poorest households (aPR=1.81 [95%CI:1.29-2.54]) compared to the not-so-poor households. Comparisons between loss of livelihood and impoverishment at the time of the interview revealed a 58% (95% CI: 52-64%) concordance. The concordance was 51% (95% 45-57%) for the period 12 months prior to the interview (**Table 4**).

**Figure 1:**
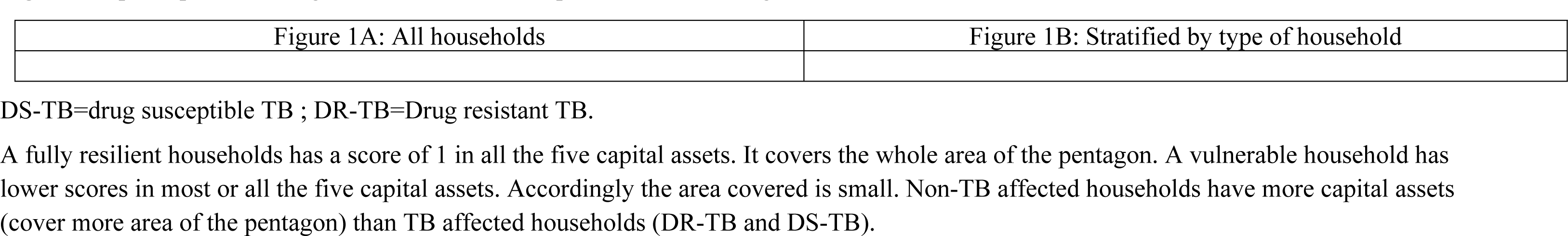
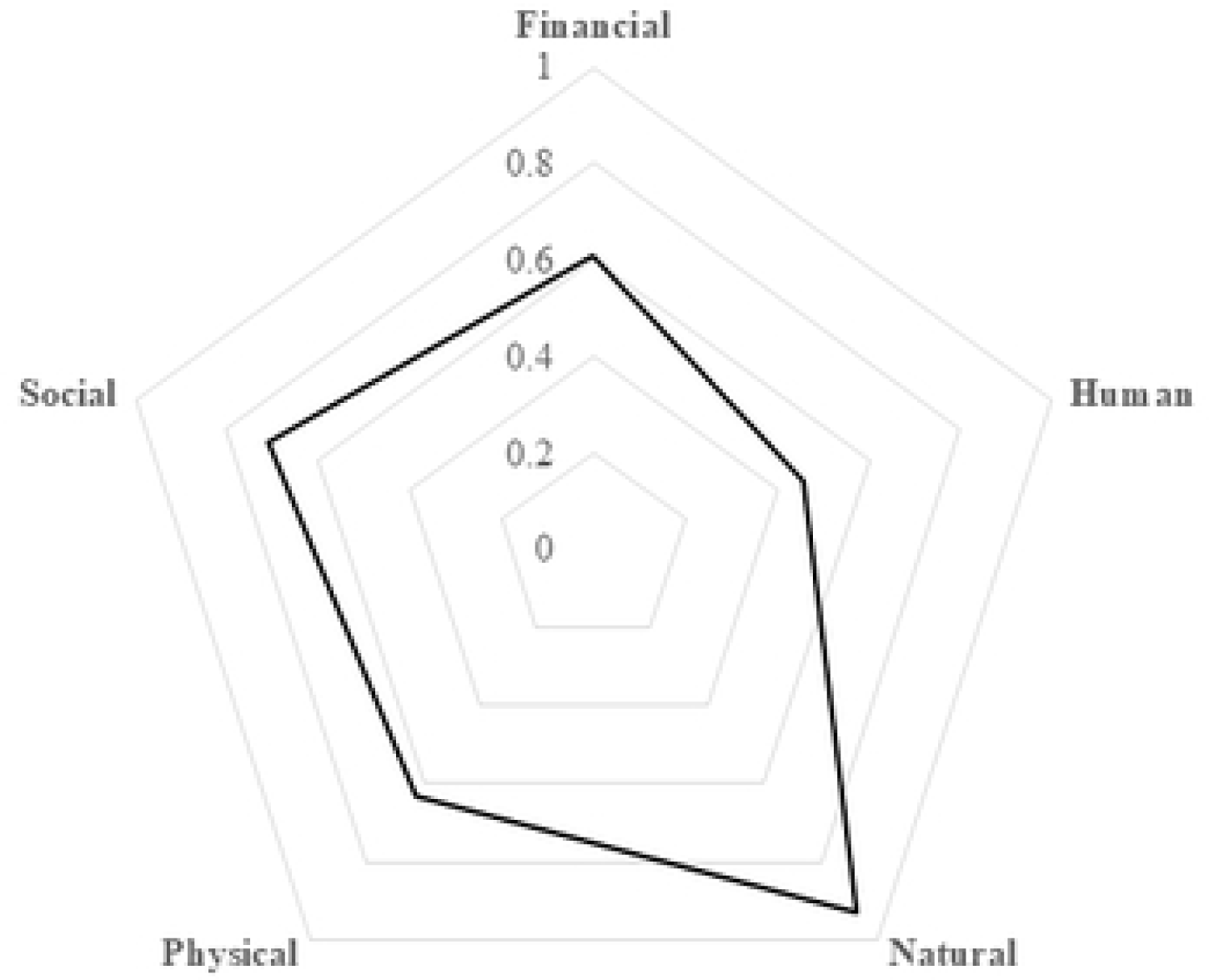
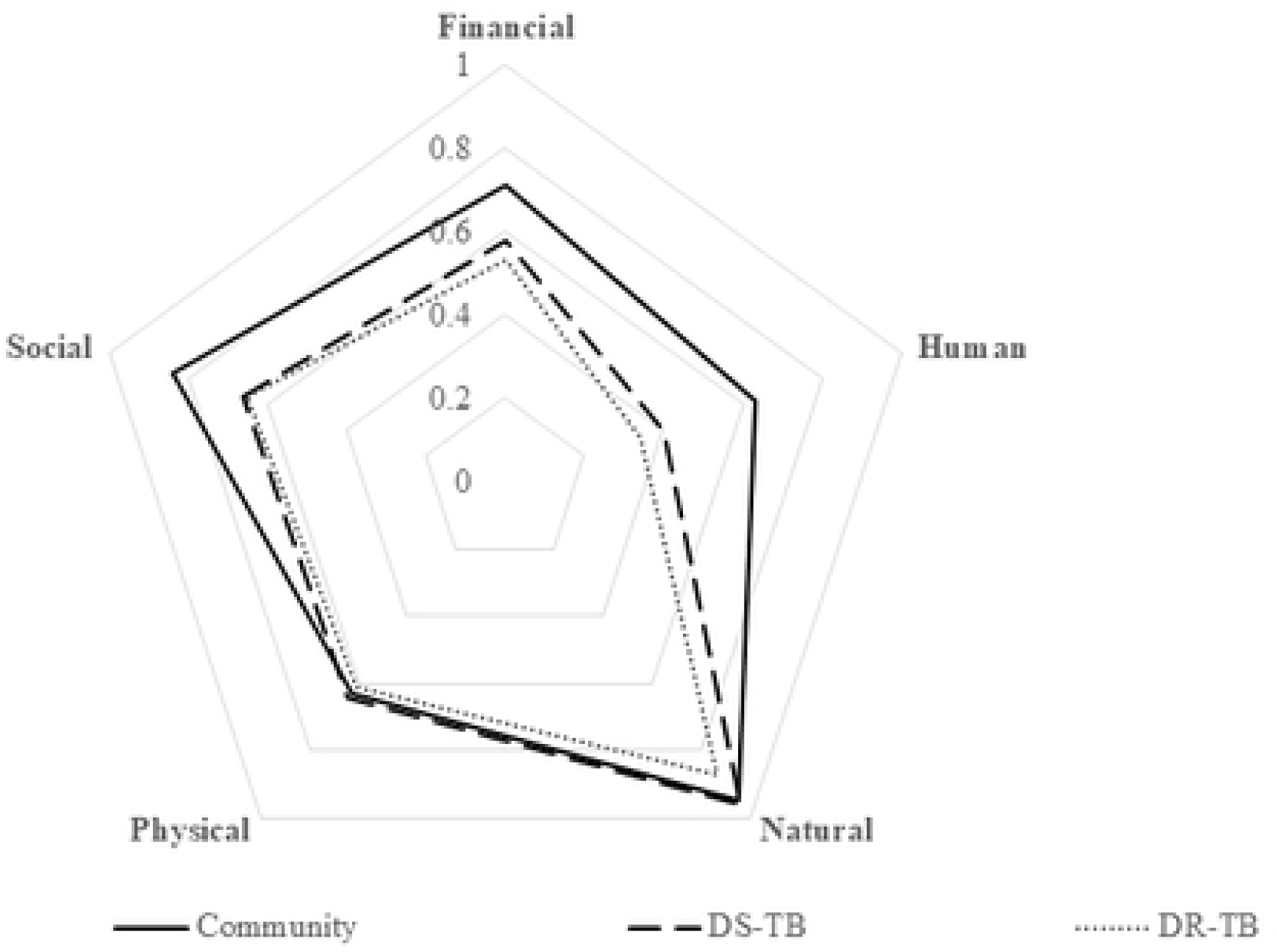
Spider-plots showing the five households capital assets following shocks such as TB and COVID-19

**Table 3:** Loss of livelihood in DS-TB, DR-TB affected households and non-TB households.

| Characteristic |  | Experienced loss of livelihood |  | PR 95% CI | aPR 95% CI |
| --- | --- | --- | --- | --- | --- |
|  |  | Total | Yes (%) * |  |  |
| Total |  | 268 | 133 (50) |  |  |
| Type of household | DR-TB | 90 | 56 (62) | 2.31 (1.58-3.37) | 2.02 (1.35-3.03) |
|  | DS-TB | 89 | 53 (60) | 2.21 (1.51-3.24) | 2.06 (1.38-3.07) |
|  | Non-TB | 89 | 24 (27) | Reference | Reference |
| History of TB in household | Yes | 109 | 57 (52) | 1.38 (1.07-1.78) | 1.06 (0.84-1.33) |
|  | No | 160 | 76(48) | Reference | Reference |
| Socioeconomic status | Poorest | 93 | 56 (60) | 1.81 (1.28-2.55) | 1.81 (1.29-2.54) |
|  | Poor | 89 | 45 (51) | 1.71 (1.19-2.44) | 1.66 (1.16-2.36) |
|  | Not so poor | 84 | 28 (33) | Reference | Reference |

\* =Row percentages; PR=prevalence ratio; aPR=adjusted prevalence ratio; CI=confidence interval; DS-TB=drug susceptible tuberculosis ; DR-TB=drug resistant tuberculosis.

**Table 4:** Concordance between impoverishment and loss of livelihood during the periods 12 months before the study and during the study.

| Impoverishment |  | Experienced loss of livelihood |  | Totals | Concordance | 95% CI |
| --- | --- | --- | --- | --- | --- | --- |
|  |  | Yes | No |  |  |  |
| 12 months before interview | Yes | 86 | 85 | 171 | 51% | (45%-57%) |
|  | No | 47 | 49 | 96 |  |  |
|  |  | 133 | 134 | 267 |  |  |
| At the time of interview | Yes | 128 | 107 | 235 | 58% | (52%-64%) |
|  | No | 5 | 27 | 32 |  |  |
|  |  | 133 | 134 | 267 |  |  |

CI=Confidence interval.

## Discussion

We used impoverishment and a multidimensional measure to investigate socioeconomic impacts of TB on households. We found that TB-affected households experience greater impoverishment and loss of livelihood than non-TB households. There was no difference in loss of livelihood between DR-TB and DS-TB affected households. Socioeconomic status was an effect modifier, and the effect of TB on loss of livelihood was worst in poorest households.

These results are in line with studies conducted in Ghana and the Philippines showing that the proportion of impoverished households is higher among TB-affected compared to non-TB households.[4], [47] Importantly, we also collected data on income 12 months prior to the interview. The proportion of TB affected households living under the poverty line 12 months prior to the interview was almost double (43%) that of non-TB affected households (23%) highlighting that TB disproportionally affects the poorest.[1] Also, there was a marked increase in impoverishment across all households from 12 months prior to the interview to the time of interview. In the 6 months prior to the interviews COVID-19 lockdowns affected businesses, public transport systems, employment opportunities, household incomes, food security and resulted in substantial impoverishment in Zimbabwe and sub-Saharan Africa.[39] Hence, impoverishment observed in this study at the time of the interview was also attributable to shocks other than TB. These finding also highlight the limitations of cross-sectional surveys which cannot account for reverse causality i.e. poor households being more likely affected by TB.

The concordance between loss of livelihood and impoverishment was 58% with the proportion impoverished being higher than the proportion which lost livelihood. This is not surprising. Impoverishment is benchmarked against income and tends to overestimate socioeconomic impact of TB among poor people, who are likely to have unstable employment or live in contexts where employment is informal and/or jobs are seasonal.(22) Income is difficult to measure, especially when the majority of the population is informally employment.[48] Furthermore, impoverishment is unable to capture the role of social support networks in mitigating socioeconomic impact of TB despite loss of income.

Most studies on socioeconomic impact of TB rely on measuring income or costs.[4], [5], [11] Our study shows that financial capital (income, spending of savings to cover TB associated costs) is not the only livelihood capital that is affected by TB. The impact of TB was found to be more pronounced on human, social and financial capitals while physical and natural capital assets remained relatively stable across all households. Natural capital is setting-specific and likely more relevant in rural areas, whilst disposal of physical capital or dilapidation of physical capital as a result of reduced maintenance may be a strategy of last resort and only employed when shocks become chronic.

Household coping strategies evolve from short term e.g. dissavings (spending savings, borrowing) to long term coping strategies (withdrawal of children from school, sale of assets). As a result, cross sectional studies, especially those in which data are collected during the intensive phase of treatment (i.e. shortly after diagnosis), may not capture long-term coping strategies. The exception may be in extremely vulnerable households, which are likely to exhaust short-term coping strategies quickly and proceed to selling assets and/or abandon treatment.[25], [49] Long-term coping strategies are the most harmful to livelihoods, with greater, long-lasting impacts. This may force households into financial catastrophes and inter-generational poverty.[50], [51] Socioeconomic impacts of TB persist even after completing treatment as households continue to borrow and pledge their assets.[7], [52], [53] Livelihood is therefore dynamic since households experience shocks continuously and are actively utilising various coping strategies in their quest to maintain well-being.[20] For this reason, longitudinal studies including the post-TB treatment period are recommended as they are likely to provide more accurate estimates of the impact of TB on households.[54]

Until recently, DS-TB and DR-TB treatment were different with regards to duration and toxicities. However, with roll out of shorter and all oral DR-TB regimens,[55], [56] the differences are less pronounced. This may explain the lack of difference in loss of livelihood between DS-TB and DR-TB households. Also, more DR-TB than DS-TB affected households reported a history of TB. People and families who previously experienced TB may be more likely to identify signs of TB early and to seek healthcare for themselves, or encourage others to do so.[10], [57] Thus it is possible that people affected by DR-TB sought care earlier, and went directly to TB clinics, resulting in fewer costs. Nevertheless, DR-TB affected households were more likely to report severe impacts of TB and other shocks on their livelihoods compared to DS-TB. This is despite no difference in the proportion experiencing loss of livelihood.

The strengths of our study include recruitment of participants across four provinces in Zimbabwe, including both urban and rural sites, and investigating socioeconomic impact of TB using a multidimensional measure which is not benchmarked against income. In parallel, we used a more conventional measure (i.e. impoverishment) allowing direct comparisons between these two. The study was undertaken during the COVID-19 pandemic, a time of extreme socioeconomic vulnerability, and provides insight into the interaction between TB and other generalised socioeconomic shocks. This has important implications for pandemic preparedness policies,[58] as it highlights the long-term impacts of pandemic responses on socioeconomic vulnerability and need to support TB-affected households especially during times of crisis.

Our cross-sectional design made it impossible to capture changes in livelihood across all phases of TB treatment. Hence, there is potential underestimation of loss of livelihood, especially among people who were interviewed during early stages of TB treatment. We relied on self-reports of coping strategies and income. While coping strategies are unlikely to be influenced by recall bias, income often is. Income is difficult to reliably estimate especially in contexts characterised by informal/seasonal jobs.[48] We potentially underestimated loss of livelihood by enrolling people who were alive and on treatment, excluding those who died of TB or were lost to follow-up prior to the study. TB-related deaths result in huge costs of up to 15 times the monthly household income.[24] People who died or were lost to follow-up are likely to have experienced greater loss of livelihood than those who were alive and on treatment. Further, we cannot rule out possible overmatching of community controls because people who live in the same area as TB-affected households often have a similar socioeconomic background. However, it is notable that the proportion of households living under the poverty line 12 months prior to the interview was significantly lower among non-TB compared to TB-affected households. Lastly, our study was conducted during the COVID-19 pandemic and both impoverishment and loss of livelihood due to TB are likely to be overestimated as concurrent shocks may have contributed to the effects seen.

Despite these limitations, our study has implications for policy and practice. Firstly, in a time of huge socioeconomic vulnerability (i.e. COVID-19), TB was associated with worse socioeconomic situation, especially in the poorest households. Hence, poorest households should be prioritised for multisectoral social protection to reduce the incidence and impacts of TB. People in the poorest households are more likely to experience food insecurity and malnutrition. Malnutrition increases risk of i) infection by *Mycobacterium tuberculosis*, ii) severe TB and iii) mortality.[59] Recent studies have shown that social protection in the form of a nutritional intervention reduces TB mortality and averts TB diseases by 40-50%.[60], [61] Secondly, this study highlights the importance of multidimensional measures to adequately capture income and non-income impacts of TB, including the effect of TB on schooling and ownership of assets, for programmatic action.[10], [62]

## Conclusion

TB affected households experienced greater loss of livelihood than households currently not affected by TB. The effect of TB is most profound among the poorest households. Multisectoral approaches to support these poorest households are crucial to mitigate the impact of TB and other shocks prevailing in communities.

## Data Availability

Data and the codebook for the study have been uploaded as supporting information.

## Conflicts of interest

None declared

## Funding statement

CT was supported by the Fogarty International Centre of the National Institutes of Health (NIH; Bethesda, MD, USA) under Award Number (D43 TW009539). The content is solely the responsibility of the authors and does not necessarily represent the official views of the National Institutes of Health. CJC (203905/Z/16/Z) and RAF (206316/Z/17/Z) are supported by the Wellcome Trust.

## Author contributions

Conceptualised the study: CT and KK

Collected data: CT, FM

Wrote the first draft: CT with support from KK, DP, VB and RH

Critically reviewed the first draft: CT, KK, DP, RH, VB, FM, CJC, RAF.

Critically reviewed final draft: CT, KK, DP, RH, VB, FM, CJC, RAF.

All authors read and approved the final manuscript.

## Data availability statement

Data supporting this manuscript can be accessed from the corresponding author upon request.

## Ethics statement

This study was approved by the Medical Research Council of Zimbabwe. All participants provided written informed consent to take part in the study.

## Acknowledgements

The authors would like to thank the participants who took part in this study. Special thanks goes to Health Services Departments (City of Harare and City of Bulawayo) as well as Provincial Medical Directorates of Masvingo and Matabeleland South.

---

**Supplement table 1:** Indicators contributing to the livelihood variable.

| Livelihood capital | Indicators |
| --- | --- |
| Financial capital | • Sale of assets.‡<br>• Spending savings.‡<br>• Failure to repay loans.<br>• Pledging future crops/cattle/livestock‡<br>• Borrowing at exorbitant interest rates‡<br>• Reduction in household income. |
| Physical capital | • Sale of productive assets.‡<br>• Failure to replace productive assets. |
| Social capital | • Changes in relationship with neighbours for the worse.<br>• Changes in relationship with family members for the worse.<br>• Received support from neighbours/family. |
| Natural capital | • Reduction in land that is farmed (idle land/leasing).<br>• Sale of land/cattle to finance TB. |
| Human capital | • Change in head of household (death/abandonment by spouse/family).<br>• Physical debility resulting in loss of labour for household.<br>• Insufficient food.<br>• Education of children affected† |

†=A derived variable obtained from either moving children to cheaper schools and/or withdrawing children from school; ‡=Coping strategies.

**Supplementary Figure 1:** Study sites.

## Supplementary file: Creating a binary variable for loss of livelihood

Use of a sustainable livelihood framework-based measure to create a dichotomous outcome variable for loss of livelihood (Yes/No).

## Household questionnaire

The household questionnaire was interviewer administered to heads of households in 180 TB affected and in 90 non-affected households. The questionnaire was administered in local language by CT (the PI) and FM (Research Assistant). The development of the questionnaire was informed by the variables in the patient cost surveys, the five capital assets on the sustainable livelihood framework, the pilot study in HIV programming in Zimbabwe, and researchers’ field experiences in Zimbabwe. Some of which were derived during analysis. Data from the paper-based questionnaires were entered in EpiData entry software for cleaning and were exported to Stata version 13.0 (StataCorp, College Station, TX, USA) for analysis.

## Variables in the household questionnaire

The outcome variable, loss of livelihood was derived from five capital assets. Variables were derived from each of the five capital assets. The variables that were included in the model are listed below.

**Table S1:** Variables included in the model to come up with an indicator for loss of livelihood.

| Capital asset | Definition | Questions | Variable name | Coding |
| --- | --- | --- | --- | --- |
| Human | Ability to work now or in the future and acquire skills | i. Was child transferred to a cheaper school? | N/A | [Yes=1/No=0] |
|  |  | ii. Was child withdrawn from school? | N/A |  |
|  |  | iii. Was education of child affected? [*derived from i and ii*] | educaffect |  |
|  |  | iv. During the past month, did you worry that your household had insufficient food? | insufood | [Yes=1/No=0] |
| Financial | Refers to cash, stock and savings. It may take the form of savings, earnings (whether regular wages or one-off payments), access to loans, or money stored in saleable property such as livestock. | i. Did your household spend savings? | spendsaving | [Yes=1/No=0] |
|  |  | ii. Did your household borrow some money | borrow | [Yes=1/No=0] |
|  |  | iii. Did your household pledge future crops or livestock? | pledged | [Yes=1/No=0] |
|  |  | iv. Did your household fail to repay loans? | failrepay | [Yes=1/No=0] |
|  |  | v. Income of household 12 months prior the interview | N/A |  |
|  |  | vi. Income of household at the time of interview | N/A |  |
|  |  | household income loss [*derived from v and vi*] | hhincomlos | [Yes=1/No=0] |
| Physical | Refers to assets such as buildings, machinery and equipment. | i. Did your household sell any assets? | soldassets | [Yes=1/No=0] |
| Social | Are relations which are based on exchange, trust and reciprocity that households depend on. | i. Relationship with family 12 months before interview on a scale of [0-10], where 0 is worst, and 10 is excellent] | N/A | N/A |
|  |  | ii. Relationship with family at the time of interview on a scale of [0-10], where 0 is worst, and 10 is excellent] | N/A | N/A |
|  |  | iii. Change in relations with family for the worse [derived variable from i and ii] | frelchange | [Yes=1/No=0] |
|  |  | iv. Relationship with neighbours 12 months before interview on a scale of [0-10], where 0 is worst, and 10 is excellent] | N/A | N/A |
|  |  | v. Relationship with neighbours at the time of interview on a scale of [0-10], where 0 is worst, and 10 is excellent] | N/A | N/A |
|  |  | vi. Change in relations with neighbours for the worse [*derived from i & ii*] | nrelchange | [Yes=1/No=0] |
|  |  | vii. Did your household receive social support (food/ cash?) | socspt | [Yes=1/No=0] |
| Natural | Include land and livestock | i. Did your household sell productive assets? | sold assets | [Yes=1/No=0] |
|  |  | ii. Did your household fail to replace the productive assets that were sold? | failrestock | [Yes=1/No=0] |

## Creating a binary variable for loss of livelihood (Yes/No) from the 13 variables

**First,** a global list of all the 13 variables was created using the syntax below. global xlist1 soldassets hhincomlos insufood spendsaving borrow failrepay prodasset educaffect pledged socspt nrelchange frelchange global id uniqueid

**Second**, the variables were correlated. Since the variables are binary (Yes/No), the tetrachoric approach was applied. The correlations were presented in a correlation matrix (**Table S1**).

### Correlations among variables

*tetrachoric $xlist1, posdef*

*adj-corr soldassets hhincomloss spendsavingg prodasset educaffect pledged failrepay insufood borrow failrestock frelchangee nrelchange socspt*

**Table S1:**
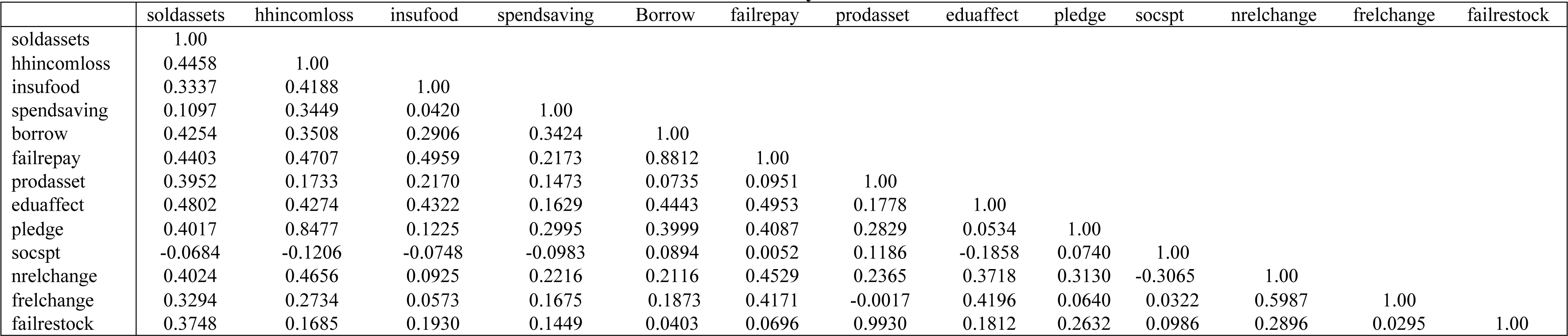
Correlation matrix of the variables included in the factor analysis.

### Measure of sampling adequacy

The Kaiser-Meyer-Olkin (KMO) was used as the measure of sampling adequacy. The following syntax was used.

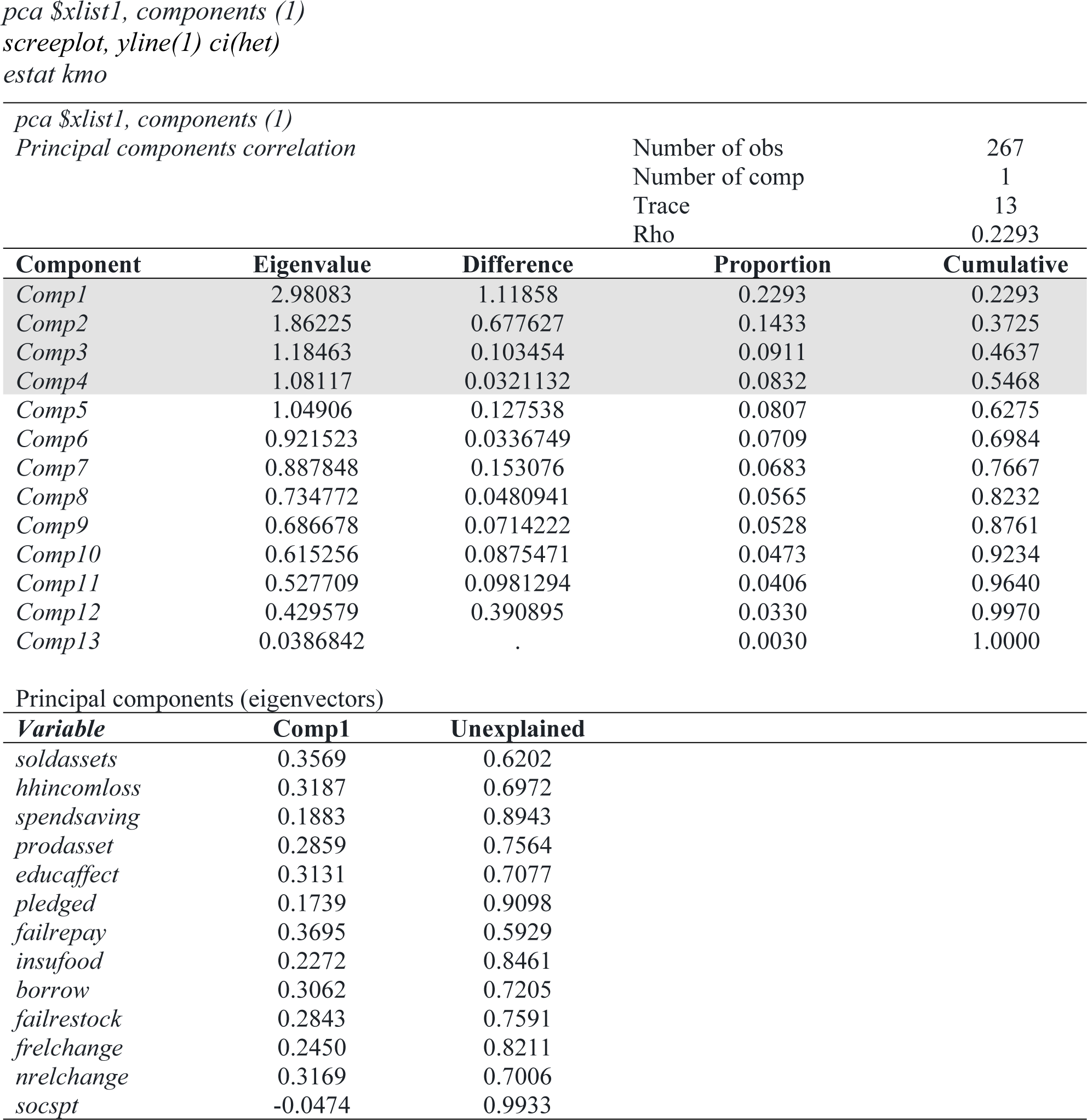

#### Screeplot

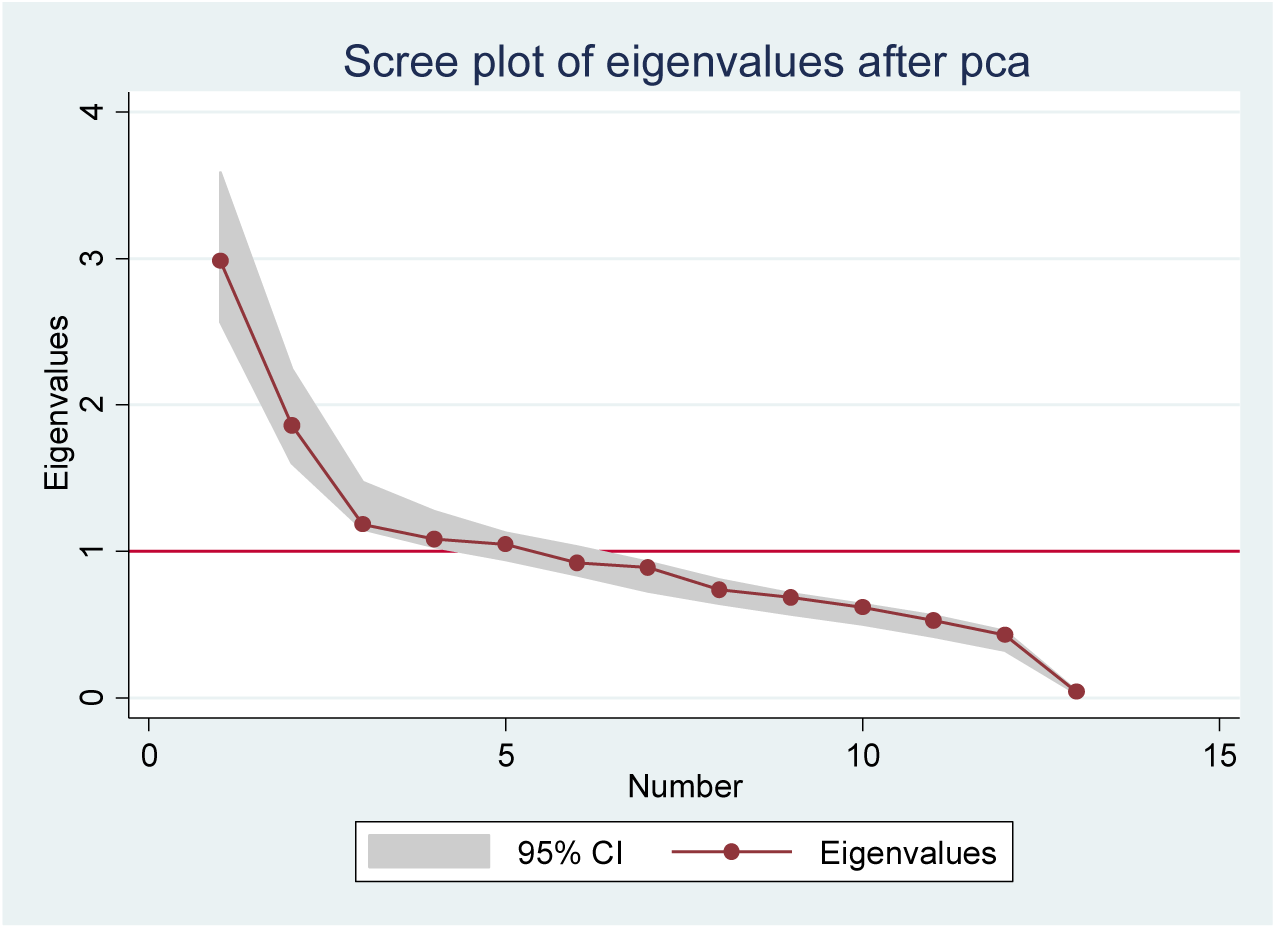

The KMO measure shows that xxxxxxxxxxxxxx

| Variable | KMO |
| --- | --- |
| soldassets | 0.8621 |
| hhincomlos | 0.7968 |
| spendsaving | 0.7098 |
| prodasset | 0.5313 |
| educaffect | 0.8401 |
| pledged | 0.7655 |
| failrepay | 0.7223 |
| insufood | 0.7395 |
| borrow | 0.6967 |
| failrestock | 0.5296 |
| frelchange | 0.6947 |
| nrelchange | 0.7017 |
| socspt | 0.4720 |
| **Overall** | **0.6588** |

| KMO measure | Interpretation |
| --- | --- |
| $KMO \geq 0.90$ | Marvellous |
| $0.80 \leq KMO < 0.90$ | Meritorious |
| $0.70 \leq KMO < 0.80$ | Average |
| $0.60 \leq KMO < 0.70$ | Mediocre |
| $0.50 \leq KMO < 0.60$ | Terrible |
| $KMO < 0.50$ | Unacceptable |

### PCA: Factor analysis, pcf

*factor $xlist1, pcf factors(1)*

### Creating binary index

*xtile lol=lol_index, nq(2)*

### Creating the index for loss of livelihood as a continuous variable

*gen lol_index =*.

*factor $xlist1, pcf factors(1)*

### Creating a Kernel density estimate to check the distribution of loss of livelihood index

*gen lol_index =*.

*factor $xlist1, pcf factors(1)*

*predict temp // SPECIFY REGRESION OR BARTLETT OPTION*

*replace lol_index = temp*

*drop temp*

*kdensity lol_index*

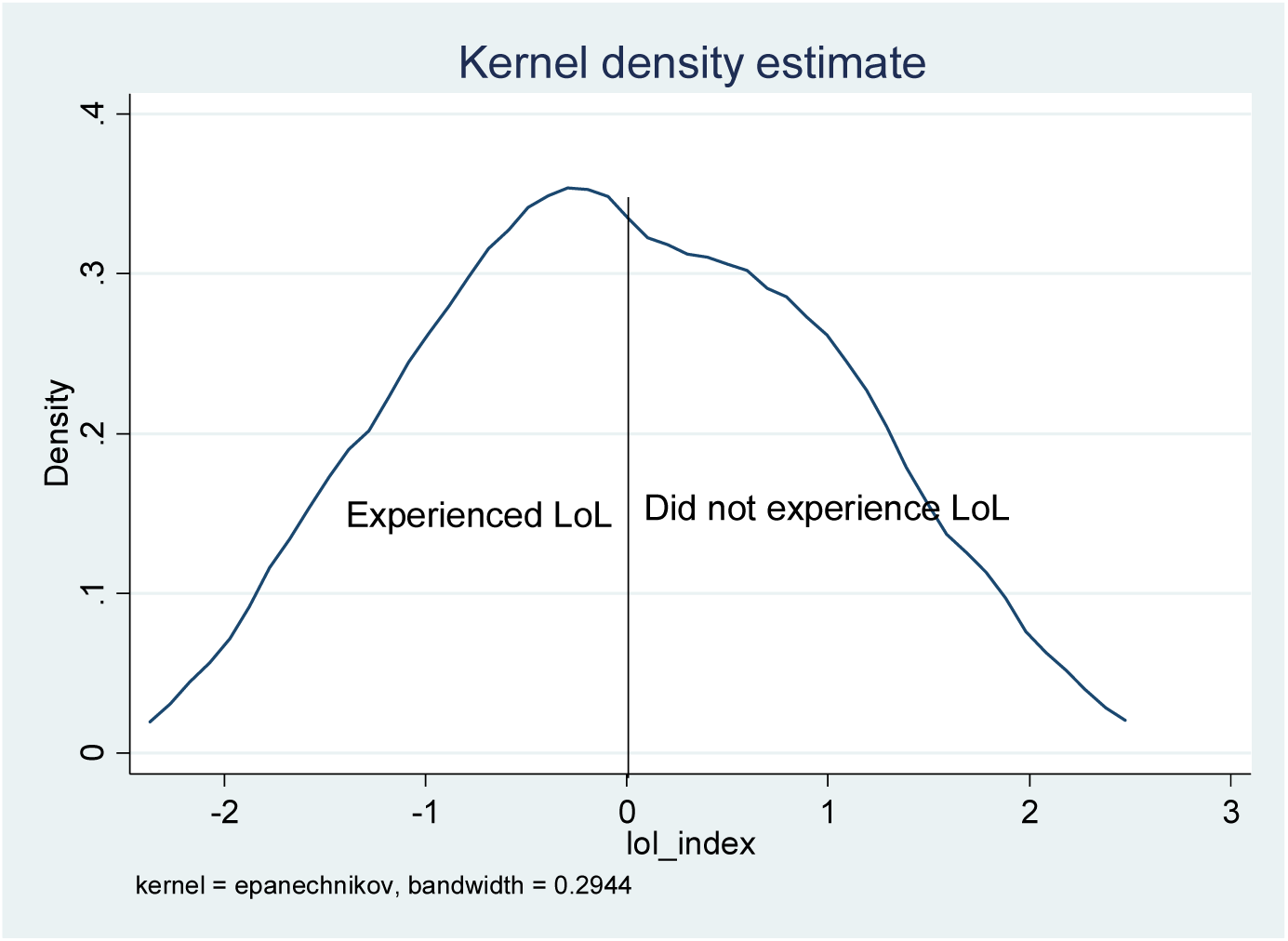

### Creating the binary variable for loss of livelihood

*gen lol=*.

*xtile temp=lol_index, nq(2)*

*replace lol = temp*

*drop temp*

*label define lol 1 “Yes” 2 “No”*

*label var lol “Loss of livelihood”*

*labelvalues lol lol*

*ta lol*

| Experienced loss of livelihood | Number | (%) |
| --- | --- | --- |
| Yes | 134 | (50.2) |
| No | 133 | (49.8) |

## Notes

### Competing Interest Statement

The authors have declared no competing interest.

